# Genetic Determinants of Chemotherapy-Induced Oral Mucositis in Children with Solid Malignancies

**DOI:** 10.64898/2026.08.05.26359776

**Authors:** Aniket Chawla, Andreas Halman, Michael See, Anneke C Grobler, Fernando J Rossello, Claire Moore, Sean Carter, Rachel Conyers

## Abstract

**Background:** Oral mucositis is a clinically significant, potentially severe side effect of systemic chemotherapy in children with cancer. Understanding genetic predisposition to this side effect may assist in development of stratified prophylactic and treatment strategies. However, existing literature primarily focuses on children with haematological malignancies.

**Methods:** We performed a candidate gene study of 101 children with solid tumours enrolled in the MARVEL-PIC study at the Royal Children’s Hospital, Melbourne. Clinical data were extracted from the electronic medical record, with NCI-CTCAE v6.0 grade ≥2 oral mucositis defined as the primary outcome. Genetic variants previously associated with oral mucositis were analysed under an additive genetic model to identify significant associations. Exploratory gene-drug interactions were identified based on chemotherapy exposure.

**Results:** 29 patients (28.7%) developed grade ≥2 oral mucositis. *MTHFR* A1298C (rs1801131) was associated with lower odds of grade ≥2 oral mucositis, lower peak mucositis grade, and lower odds of opioid use for oral mucositis. 25 exploratory gene-drug interaction signals were identified, including miR-1206 rs2114358 with methotrexate exposure and *ABCB1* rs1045642 with anthracycline exposure.

**Conclusions:** *MTHFR* A1298C (rs1801131) demonstrated a protective effect against chemotherapy-induced oral mucositis in our cohort of children with solid tumours. Larger, ancestry-informed studies are required to validate our findings.

## 1.0 Background

Oral mucositis, defined as inflammation of the oral mucosa, is a common toxicity of chemotherapy in children with cancer. Its reported incidence varies from 52-100% and is higher among patients receiving high-dose chemotherapy.^1^ In a cohort of 111 patients with Ewing sarcoma, 39.1% developed at least one episode of NCI-CTCAE (National Cancer Institute Common Terminology Criteria for Adverse Events) grade ≥3 oral mucositis during induction chemotherapy.^2^ Oral mucositis can cause severe pain, impaired oral intake, and subsequent malnutrition, greatly reducing a child’s quality of life.^3^ Qualitative literature has highlighted severe psychosocial impacts of oral mucositis on children, including communication difficulties, complex relationships with food, and deviations from a sense of “normality”.^4^ Oral mucositis may also disrupt cancer treatment; a 2024 retrospective review of 200 patients identified that 110 (55%) required dose reduction (n=66; 33%), delay (n=42; 21%), or discontinuation (n=2; 1%) of therapy.^5^ Oral mucositis is also associated with a substantial economic burden; a 2021 systematic review reported that its cost reached up to $31963.64 USD per patient in some treatment settings.^6^

The pathophysiology of oral mucositis comprises five stages: (i) initiation of mucosal injury, (ii) primary damage response, (iii) signal amplification, (iv) ulceration, and (v) healing.^7^ Chemotherapy drugs that are commonly implicated in mucositis include antimetabolites (e.g. methotrexate, fluorouracil), alkylating agents (e.g. cyclophosphamide, ifosfamide), platinum agents (e.g. cisplatin, carboplatin), and anthracyclines (e.g. doxorubicin, daunorubicin).^8^ Management is primarily supportive care (basic oral care, anti-inflammatory medications, analgesia), and may also include antimicrobials, photobiomodulation, and nutritional support.^9^

Inter-individual genetic variation may influence both the pathways involved in mucosal injury and repair, as well as the metabolism and transport of chemotherapy drugs, potentially increasing drug accumulation and risk of toxicity. Several genetic variants have previously been associated with chemotherapy-induced oral mucositis, including in *MTHFR*, *ABCB1*, *ABCG2* and *SLCO1B1*.^10^ However, evidence to support the role of these variants is inconsistent, warranting further investigation. *MTHFR* C677T (rs1801133), which makes the MTHFR (methylenetetrahydrofolate reductase) enzyme more thermolabile, thereby decreasing its activity, is among the most extensively studied genetic variant.^11^ Reduced MTHFR activity may alter folate availability and impair nucleotide synthesis for rapidly-dividing mucosal cells, increasing susceptibility to chemotherapy-induced mucosal injury. However, previous studies investigating this polymorphism have yielded inconsistent findings.^10^ *ABCB1* C3435T (rs1045642) has also been investigated in regard to its association with oral mucositis. *ABCB1* encodes the ATP-binding cassette of subfamily B, which functions as a drug efflux pump.^12^ The rs1045642 polymorphism has been associated with reduced transporter activity and thus may lead to prolonged exposure of rapidly-dividing mucosal cells to cytotoxic drugs, increasing a patient’s risk of toxicity.

Notably, the current literature on genetic determinants of chemotherapy-induced oral mucositis in children predominantly focuses on children with haematological malignancies. Evidence focussed on genetic predisposition to mucositis in patients with a solid malignancy is limited and primarily focuses on single diseases such as Ewing sarcoma or osteosarcoma.^2, 13, 14^ We therefore aimed to evaluate whether any selected candidate genetic variants are associated with the incidence and severity of chemotherapy-induced oral mucositis in children with solid malignancies. We also examined clinical factors, including age and sex, as potential contributors to mucositis risk

## 2.0 Methods

This study was a sub-study of the MARVEL-PIC randomised controlled trial (NCT05667766) approved by the Royal Children’s Hospital Ethics Committee (HREC/89083/RCHM-2022). Ethical approval for this sub-study was obtained from the Research Ethics & Governance office of the Royal Children’s Hospital, Melbourne (REG 4868).

### 2.1 Patient Selection

Patients enrolled in the MARVEL-PIC trial with available genetic data were considered for inclusion.^15^ Children aged 0–18 years with a first cancer diagnosis of a solid malignancy subtype receiving their first two cycles of chemotherapy at the Royal Children’s Hospital were included. Exclusion criteria included secondary malignancy or relapse of primary disease, patients who did not complete two cycles of chemotherapy or received their first two cycles at other institutions, CNS tumours, and patients with incomplete clinical and/or genetic data.

### 2.2 Clinical Data

Clinical data were extracted from the electronic medical record. Extracted data included demographics (age at first cycle of chemotherapy and sex at birth), cancer diagnosis, treatment (drugs received during first two cycles, chemotherapy regimens), mucositis outcomes (incidence, severity), and additional indicators of oral mucositis severity (opioid use, total parenteral nutrition, and unplanned admissions attributed to oral mucositis). Self-reported ancestry data were collected from the MARVEL-PIC database; patients were given the option to select from seven broad pre-colonisation biogeographical groups, and their selections were reported. Oral mucositis was retrospectively graded based on available clinical documentation using NCI-CTCAE version 6.0, with grade ≥2 oral mucositis defined as the primary outcome.^16^

### 2.3 Genetic Data

Genetic variants previously implicated in chemotherapy-induced oral mucositis in children were identified through a targeted literature search. Variants with any previously reported association were selected for analysis (Table 1). *MTHFR* rs1801133 (c.677C>T) and *ABCB1* rs1045642 (c.3435C>T) were considered primary variants of interest, as they have previously displayed significant associations with oral mucositis in multiple cohorts.^10, 17^

**Table 1:**
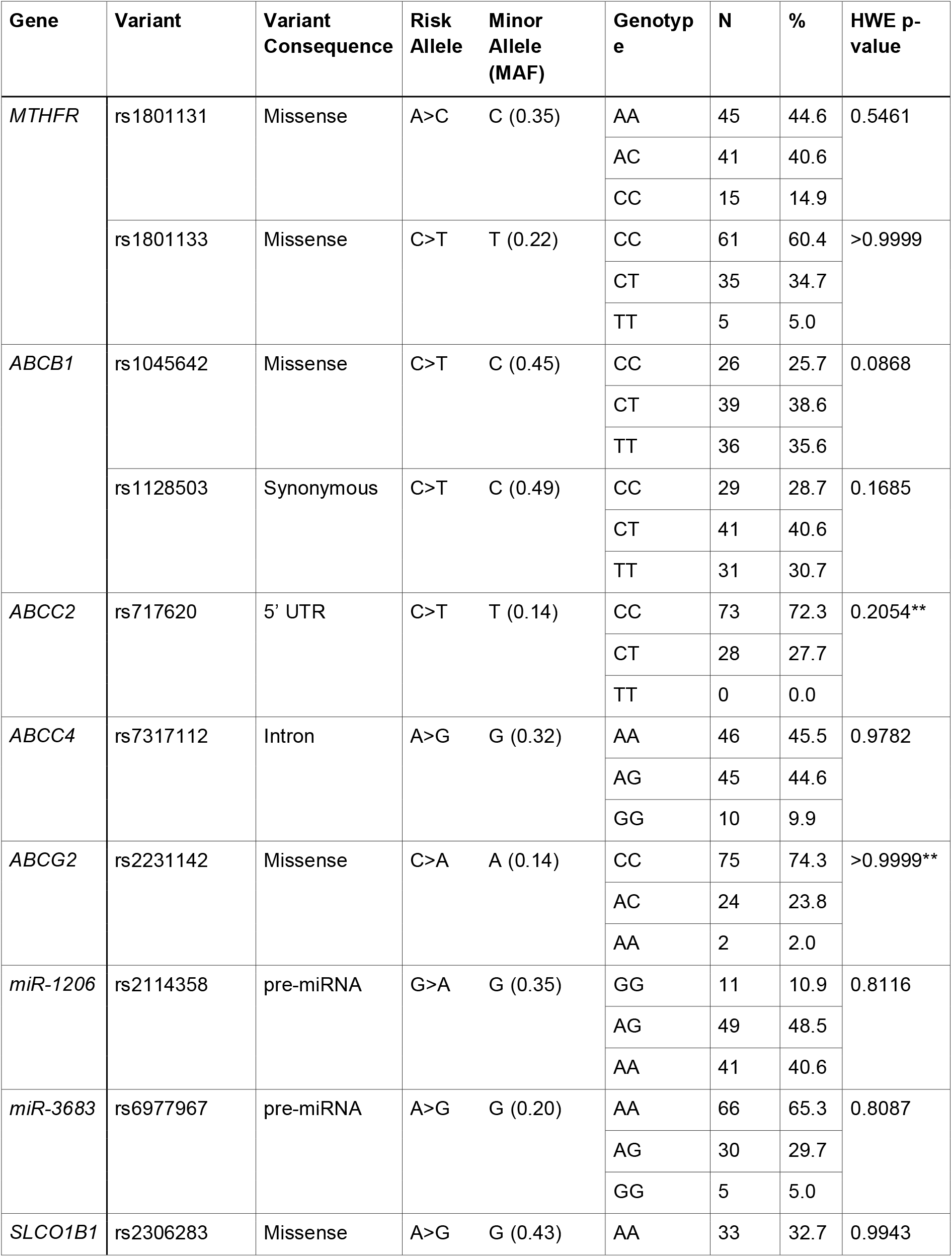
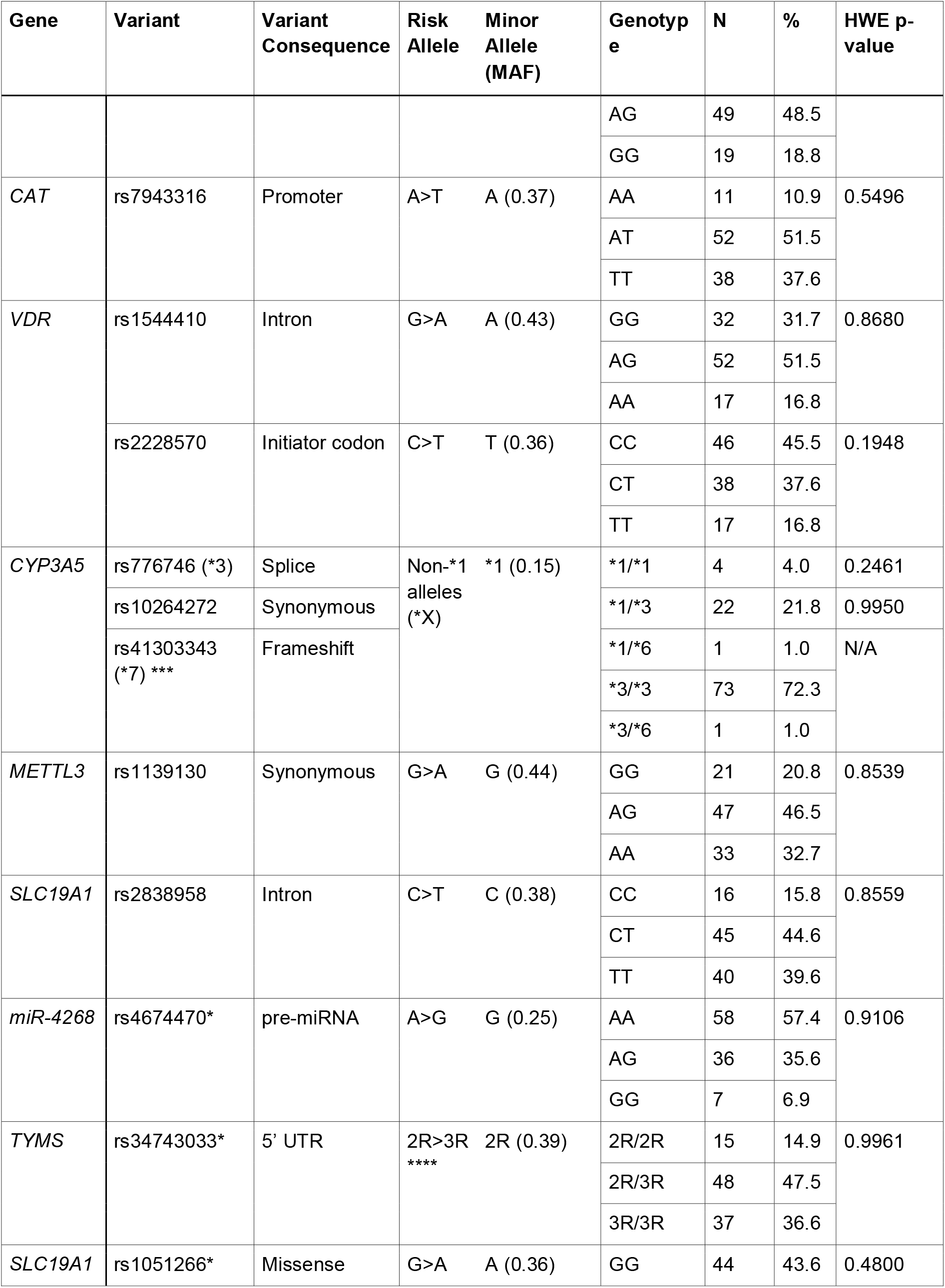
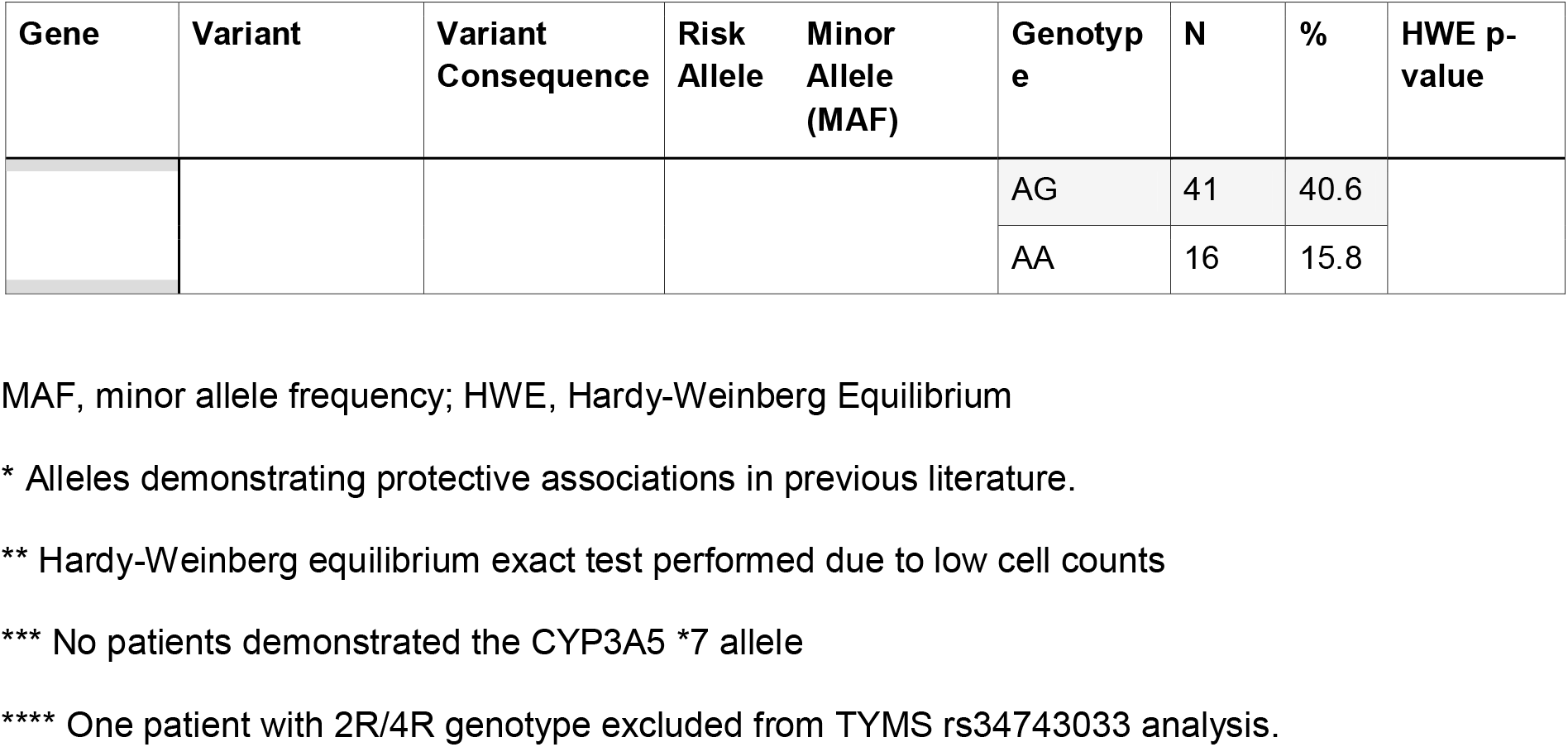
Variants investigated in present study.

| Gene | Variant | Variant Consequence | Risk Allele | Minor Allele (MAF) | Genotype | N | % | HWE p-value |
| --- | --- | --- | --- | --- | --- | --- | --- | --- |
| *MTHFR* | rs1801131 | Missense | A>C | C (0.35) | AA | 45 | 44.6 | 0.5461 |
|  |  |  |  |  | AC | 41 | 40.6 |  |
|  |  |  |  |  | CC | 15 | 14.9 |  |
|  | rs1801133 | Missense | C>T | T (0.22) | CC | 61 | 60.4 | >0.9999 |
|  |  |  |  |  | CT | 35 | 34.7 |  |
|  |  |  |  |  | TT | 5 | 5.0 |  |
| *ABCB1* | rs1045642 | Missense | C>T | C (0.45) | CC | 26 | 25.7 | 0.0868 |
|  |  |  |  |  | CT | 39 | 38.6 |  |
|  |  |  |  |  | TT | 36 | 35.6 |  |
|  | rs1128503 | Synonymous | C>T | C (0.49) | CC | 29 | 28.7 | 0.1685 |
|  |  |  |  |  | CT | 41 | 40.6 |  |
|  |  |  |  |  | TT | 31 | 30.7 |  |
| *ABCC2* | rs717620 | 5' UTR | C>T | T (0.14) | CC | 73 | 72.3 | 0.2054** |
|  |  |  |  |  | CT | 28 | 27.7 |  |
|  |  |  |  |  | TT | 0 | 0.0 |  |
| *ABCC4* | rs7317112 | Intron | A>G | G (0.32) | AA | 46 | 45.5 | 0.9782 |
|  |  |  |  |  | AG | 45 | 44.6 |  |
|  |  |  |  |  | GG | 10 | 9.9 |  |
| *ABCG2* | rs2231142 | Missense | C>A | A (0.14) | CC | 75 | 74.3 | >0.9999** |
|  |  |  |  |  | AC | 24 | 23.8 |  |
|  |  |  |  |  | AA | 2 | 2.0 |  |
| *miR-1206* | rs2114358 | pre-miRNA | G>A | G (0.35) | GG | 11 | 10.9 | 0.8116 |
|  |  |  |  |  | AG | 49 | 48.5 |  |
|  |  |  |  |  | AA | 41 | 40.6 |  |
| *miR-3683* | rs6977967 | pre-miRNA | A>G | G (0.20) | AA | 66 | 65.3 | 0.8087 |
|  |  |  |  |  | AG | 30 | 29.7 |  |
|  |  |  |  |  | GG | 5 | 5.0 |  |
| *SLCO1B1* | rs2306283 | Missense | A>G | G (0.43) | AA | 33 | 32.7 | 0.9943 |
|  |  |  |  |  | AG | 49 | 48.5 |  |
|  |  |  |  |  | GG | 19 | 18.8 |  |
| CAT | rs7943316 | Promoter | A>T | A (0.37) | AA | 11 | 10.9 | 0.5496 |
|  |  |  |  |  | AT | 52 | 51.5 |  |
|  |  |  |  |  | TT | 38 | 37.6 |  |
| VDR | rs1544410 | Intron | G>A | A (0.43) | GG | 32 | 31.7 | 0.8680 |
|  |  |  |  |  | AG | 52 | 51.5 |  |
|  |  |  |  |  | AA | 17 | 16.8 |  |
|  | rs2228570 | Initiator codon | C>T | T (0.36) | CC | 46 | 45.5 | 0.1948 |
|  |  |  |  |  | CT | 38 | 37.6 |  |
|  |  |  |  |  | TT | 17 | 16.8 |  |
| CYP3A5 | rs776746 (*3) | Splice | Non-*1 alleles (*X) | *1 (0.15) | *1/*1 | 4 | 4.0 | 0.2461 |
|  | rs10264272 | Synonymous |  |  | *1/*3 | 22 | 21.8 | 0.9950 |
|  | rs41303343 (*7) *** | Frameshift |  |  | *1/*6 | 1 | 1.0 | N/A |
|  |  |  |  |  | *3/*3 | 73 | 72.3 |  |
|  |  |  |  |  | *3/*6 | 1 | 1.0 |  |
| METTL3 | rs1139130 | Synonymous | G>A | G (0.44) | GG | 21 | 20.8 | 0.8539 |
|  |  |  |  |  | AG | 47 | 46.5 |  |
|  |  |  |  |  | AA | 33 | 32.7 |  |
| SLC19A1 | rs2838958 | Intron | C>T | C (0.38) | CC | 16 | 15.8 | 0.8559 |
|  |  |  |  |  | CT | 45 | 44.6 |  |
|  |  |  |  |  | TT | 40 | 39.6 |  |
| miR-4268 | rs4674470* | pre-miRNA | A>G | G (0.25) | AA | 58 | 57.4 | 0.9106 |
|  |  |  |  |  | AG | 36 | 35.6 |  |
|  |  |  |  |  | GG | 7 | 6.9 |  |
| TYMS | rs34743033* | 5' UTR | 2R>3R **** | 2R (0.39) | 2R/2R | 15 | 14.9 | 0.9961 |
|  |  |  |  |  | 2R/3R | 48 | 47.5 |  |
|  |  |  |  |  | 3R/3R | 37 | 36.6 |  |
| SLC19A1 | rs1051266* | Missense | G>A | A (0.36) | GG | 44 | 43.6 | 0.4800 |
|  |  |  |  |  | AG | 41 | 40.6 |  |
|  |  |  |  |  | AA | 16 | 15.8 |  |

MAF, minor allele frequency; HWE, Hardy-Weinberg Equilibrium
\* Alleles demonstrating protective associations in previous literature.
\*\* Hardy-Weinberg equilibrium exact test performed due to low cell counts
\*\*\* No patients demonstrated the CYP3A5 \*7 allele
\*\*\*\* One patient with 2R/4R genotype excluded from TYMS rs34743033 analysis.

Whole genome sequencing (WGS) was performed by the Victorian Clinical Genetics Services (VCGS) or the Australian Genome Research Facility (AGRF). DNA was extracted using the QIAsymphony DNA Mini Kit, libraries were prepared using the Illumina DNA PCR-Free Prep workflow, and sequencing was performed on the Illumina NovaSeq X platform, targeting a minimum sequencing depth of 27x and generating paired-end 150 bp reads. Raw sequencing data were processed using a clinically accredited alignment and variant-calling pipeline at VCGS. Reads were aligned to the GRCh38 reference genome using DRAGEN v4.0.5 (Illumina), and variants were called using the DRAGEN Germline Small Variant Caller. Variants of interest were extracted from the variant call format (VCF) file using bcftools v1.19 and a BED file (**Supplementary Material S1**) containing the relevant genetic variants. Regions containing variants that were not called in any samples were manually reviewed to confirm read coverage. Sequencing and variant calling were performed prior to clinical data collection and thus were independent of mucositis outcome. Genetic data were collected in small batches of samples, though with the same methodology (DNA extraction, sequencing, variant calling, and variant extraction).

### 2.4 Data Analysis

An a priori power calculation was performed for the primary pharmacogenomic analysis. Assuming an additive genetic model, a minor allele frequency (MAF) of 0.30, grade ≥2 mucositis in 40% of participants and two-sided α = 0.05, this study had 35% statistical power to detect a moderate genetic effect (odds ratio = 2.0) for the primary variants of interest.

Summary statistics of sex, age, ancestral group, diagnosis, chemotherapy exposures, and mucositis outcomes were calculated using Microsoft Excel. Associations of age and sex with the incidence of oral mucositis (grade ≥2) were analysed using univariate logistic regression.

Genotypes were coded using an additive genetic model, with each individual assigned a value of 0, 1, or 2 according to the number of risk alleles present for each genotype. *CYP3A5* genotype was coded according to the number of non-*1 alleles present. *TYMS* genotype was coded according to the number of 3R alleles present, with genotypes 2R/2R, 2R/3R, and 3R/3R assigned values of 0, 1, and 2, respectively. One participant with genotype 2R/4R was excluded from the *TYMS* analysis, as the 4R allele was rare in our cohort (frequency 0.5%). Hardy-Weinberg equilibrium (HWE) was assessed for each genetic variant using the chi-squared test or Hardy-Weinberg exact tests if expected cell counts were less than five.^18^

For primary analysis, logistic regression was used to estimate the association between primary variants of interest (*MTHFR* rs1801133 and *ABCB1* rs1045642) and the incidence of NCI-CTCAE grade ≥2 oral mucositis, as well as other indicators of mucositis severity including opioid use, total parenteral nutrition (TPN), and unplanned admissions attributable to oral mucositis. Associations between the remaining genetic variants and grade ≥2 oral mucositis, as well as severity indicators of oral mucositis, were also assessed using logistic regression, with false discovery rate (FDR) controlled using the Benjamini-Hochberg method (q = 0.10).^19^ Models that failed to converge due to sparse genotype or outcome counts were reported as not estimable and were excluded from Benjamini–Hochberg FDR correction. Ordinal logistic regression was used to assess the association between each genetic variant and peak oral mucositis grade.

Sensitivity analyses were performed in two ways. First, alternative mucositis outcome thresholds of grade ≥1 and grade ≥3 were assessed under the additive genetic model. Second, associations with grade ≥2 oral mucositis were assessed under dominant and recessive genetic models. Benjamini–Hochberg FDR correction was used for sensitivity analyses as described above.

Separate multivariable logistic regression models were fitted to assess interactions between genotype and exposure to platinum compounds (cisplatin or carboplatin), alkylating agents (cyclophosphamide or ifosfamide), anthracyclines (doxorubicin or daunorubicin), etoposide, methotrexate, and actinomycin D in relation to grade ≥2 oral mucositis. An interaction term between drug exposure and genotype was included in each model, with a p-value <0.2 used to identify exploratory gene-drug interactions.

P-values <0.05 were considered statistically significant where Benjamini-Hochberg correction for FDR was not used. For gene-drug interactions, p<0.2 was considered suggestive. GraphPad Prism was used for all statistical analyses except ordinal logistic regression, which was performed using jamovi.

## 3.0 Results

123 patients with solid malignancies enrolled in the MARVEL-PIC (n=441) study at the Royal Children’s Hospital, Melbourne were identified. Of these, 101 were identified for inclusion (**Figure 1**).

Of 101 included patients, 57 (56.4%) were male and 44 (43.6%) were female. Median age at first chemotherapy cycle was 5.3 years (range 0.0–17.3 years). Self-reported ancestry groups included European (n=56; 55.4%), Oceanian (n=29; 28.7%), Central/South Asian (n=12; 11.9%), East Asian (n=10; 9.9%), American (n=5; 5.0%), Near Eastern (n=5; 5.0%), and Sub-Saharan African (n=4; 4.0%). 65 (64.4%) identified with one ancestral group, 27 (26.7%) with two groups, and 2 (2.0%) with three groups. Seven patients did not have self-reported ancestry data available through MARVEL-PIC. The most common diagnoses were Hodgkin Lymphoma (n=15; 14.9%), rhabdomyosarcoma (n=13; 12.9%), Ewing sarcoma (n=12; 11.9%), osteosarcoma (n=12; 11.9%), and neuroblastoma (n=11; 10.9%). The most administered mucotoxic drugs were doxorubicin (n=65; 64.4%), etoposide (n=42; 41.6%), cyclophosphamide (n=31; 30.7%), and ifosfamide (n=30; 29.7%).

29 patients (28.7%) developed NCI-CTCAE grade ≥2 oral mucositis. 25 patients (24.8%) were prescribed opioids for oral pain, 5 (5.0%) required TPN, and 28 (27.7%) had unplanned admissions secondary to their oral mucositis. Clinical characteristics and outcomes are described in **Table 2**. Neither age nor sex were associated with grade ≥2 oral mucositis, but age was associated with TPN requirement for oral mucositis (OR 0.62 [0.25–0.91], p=0.0046). Sensitivity analyses also indicated increasing age was associated with grade ≥1 oral mucositis (OR 1.083 [1.010–1.165], p = 0.0245).

**Table 2:**
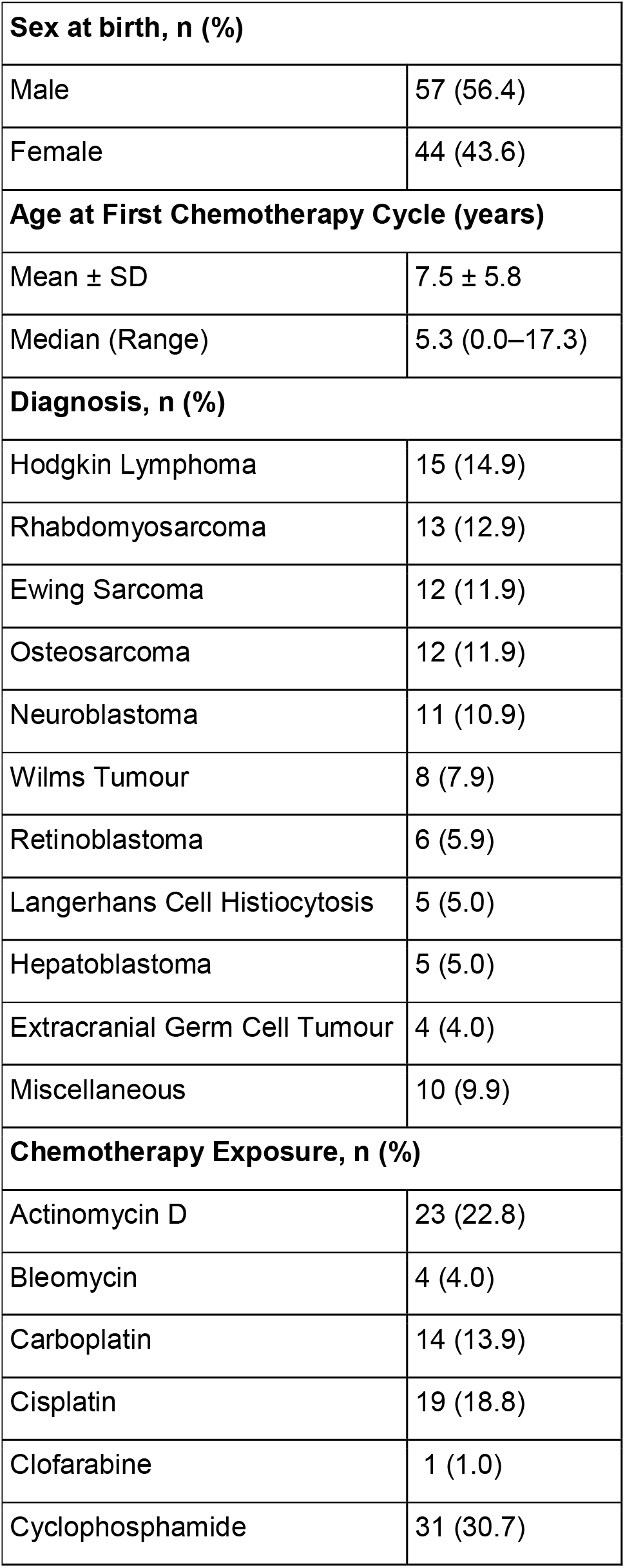
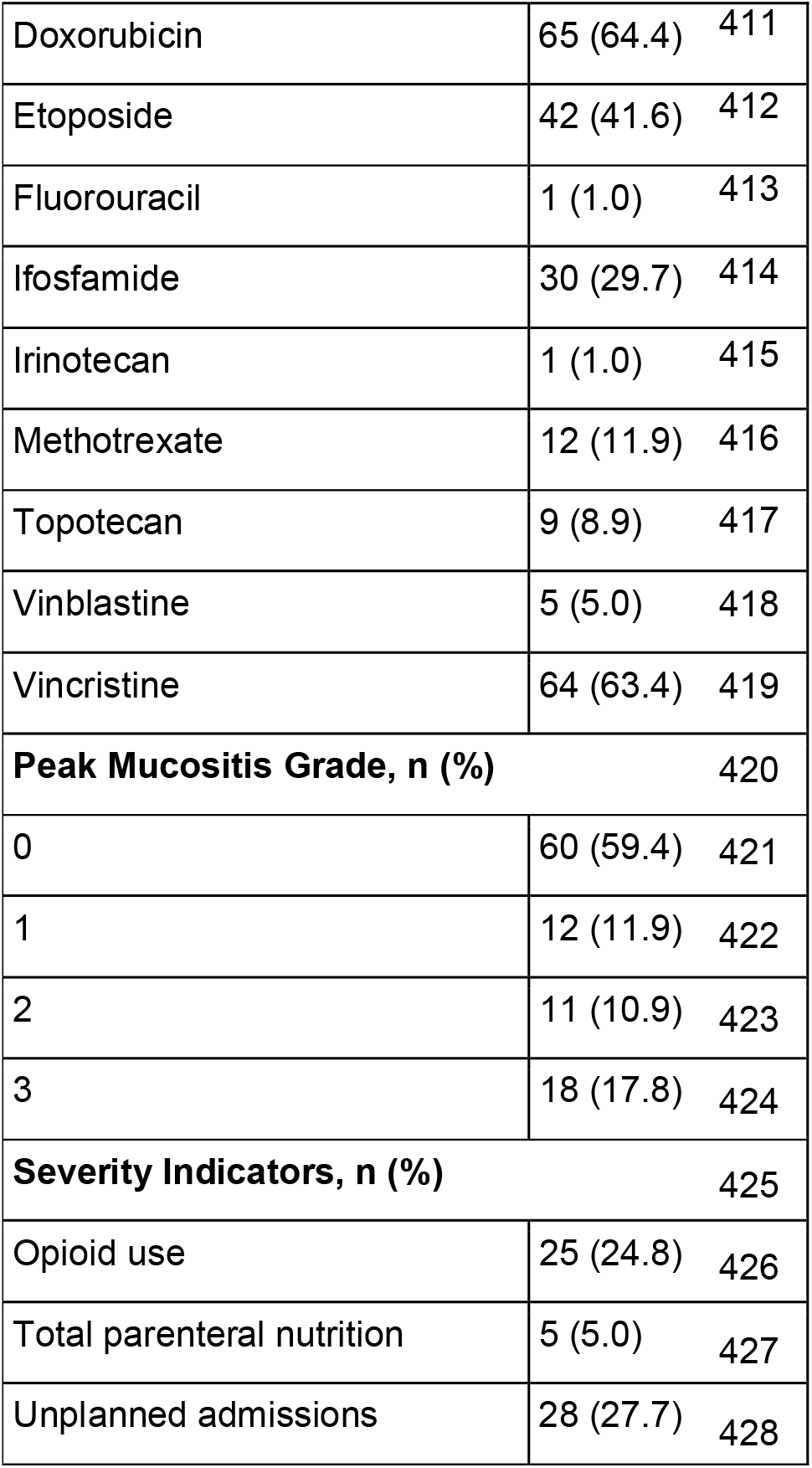
Clinical characteristics of study population.

|  |  |
| --- | --- |
| **Sex at birth, n (%)** |  |
| Male | 57 (56.4) |
| Female | 44 (43.6) |
| **Age at First Chemotherapy Cycle (years)** |  |
| Mean ± SD | 7.5 ± 5.8 |
| Median (Range) | 5.3 (0.0–17.3) |
| **Diagnosis, n (%)** |  |
| Hodgkin Lymphoma | 15 (14.9) |
| Rhabdomyosarcoma | 13 (12.9) |
| Ewing Sarcoma | 12 (11.9) |
| Osteosarcoma | 12 (11.9) |
| Neuroblastoma | 11 (10.9) |
| Wilms Tumour | 8 (7.9) |
| Retinoblastoma | 6 (5.9) |
| Langerhans Cell Histiocytosis | 5 (5.0) |
| Hepatoblastoma | 5 (5.0) |
| Extracranial Germ Cell Tumour | 4 (4.0) |
| Miscellaneous | 10 (9.9) |
| **Chemotherapy Exposure, n (%)** |  |
| Actinomycin D | 23 (22.8) |
| Bleomycin | 4 (4.0) |
| Carboplatin | 14 (13.9) |
| Cisplatin | 19 (18.8) |
| Clofarabine | 1 (1.0) |
| Cyclophosphamide | 31 (30.7) |

|  |  |  |
| --- | --- | --- |
| Doxorubicin | 65 (64.4) | 411 |
| Etoposide | 42 (41.6) | 412 |
| Fluorouracil | 1 (1.0) | 413 |
| Ifosfamide | 30 (29.7) | 414 |
| Irinotecan | 1 (1.0) | 415 |
| Methotrexate | 12 (11.9) | 416 |
| Topotecan | 9 (8.9) | 417 |
| Vinblastine | 5 (5.0) | 418 |
| Vincristine | 64 (63.4) | 419 |
| **Peak Mucositis Grade, n (%)** |  | 420 |
| 0 | 60 (59.4) | 421 |
| 1 | 12 (11.9) | 422 |
| 2 | 11 (10.9) | 423 |
| 3 | 18 (17.8) | 424 |
| **Severity Indicators, n (%)** |  | 425 |
| Opioid use | 25 (24.8) | 426 |
| Total parenteral nutrition | 5 (5.0) | 427 |
| Unplanned admissions | 28 (27.7) | 428 |

Minor allele frequency within our cohort ranged from 0.14 (*ABCC2* rs717620 and *ABCG2* rs2231142) to 0.49 (*ABCB1* rs1128503) (**Table 1**). No genetic variants deviated significantly from Hardy-Weinberg Equilibrium. One variant (*CYP3A5* *7) was not identified in any patients. One patient demonstrated genotype 2R/4R for *TYMS* rs34743033 and was excluded from further analysis for this genetic variant.

Univariate logistic regression demonstrated a significant protective association between *MTHFR* A1298C (rs1801131) and NCI-CTCAE grade ≥2 mucositis (OR 0.35 [0.16–0.70], p=0.0024) (**Table 3**). This remained significant after Benjamini-Hochberg FDR correction (significance level p<0.0059). No other investigated variants demonstrated significant associations with incidence of grade ≥2 oral mucositis. Sensitivity analyses under a dominant gene model retained this significant association, but a logistic regression model could not be fitted under a recessive model (**Supplementary Table S2**). Ordinal logistic regression also indicated a significant association between this variant and lower peak mucositis grade (p<0.001, FDR-corrected significance level p<0.006); no other variants reached significance (**Supplementary Table S3**).

**Table 3:** Association of Genetic Variants with Grade ≥2 Mucositis (Logistic Regression)

| Variant | OR [95% CI] | p-value |
| --- | --- | --- |
| *MTHFR* rs1801131 | 0.35 [0.16–0.70] | 0.0024** |
| *MTHFR* rs1801133* | 1.52 [0.74–3.11] | 0.2557 |
| *ABCB1* rs1045642* | 1.10 [0.63–1.93] | 0.7492 |
| *ABCB1* rs1128503 | 0.88 [0.50–1.54] | 0.6531 |
| *ABCC2* rs717620 | 1.58 [0.61–3.99] | 0.3415 |
| *ABCC4* rs7317112 | 0.83 [0.41–1.60] | 0.5737 |
| *ABCG2* rs2231142 | 0.64 [0.22–1.58] | 0.3474 |
| miR-1206 rs2114358 | 0.75 [0.38–1.44] | 0.3805 |
| miR-3683 rs6977967 | 1.41 [0.68–2.90] | 0.3483 |
| *SLCO1B1* rs2306283 | 0.91 [0.49–1.68] | 0.7592 |
| *CAT* rs7943316 | 0.86 [0.43–1.69] | 0.6689 |
| *VDR* rs1544410 | 0.87 [0.46–1.65] | 0.6730 |
| *VDR* rs2228570 | 1.46 [0.82–2.63] | 0.1989 |
| *CYP3A5* (non-*1 alleles) | 0.72 [0.34–1.59] | 0.4010 |
| *METTL3* rs1139130 | 0.73 [0.39–1.32] | 0.2939 |
| *SLC19A1* rs2838958 | 1.12 [0.61–2.10] | 0.7290 |
| miR-4268 rs4674470 | 0.74 [0.34–1.48] | 0.3994 |
| *TYMS* rs34743033 | 0.96 [0.51–1.82] | 0.9030 |
| *SLC19A1* rs1051266 | 1.10 [0.60–2.00] | 0.7510 |

\* primary variants
\*\* significant ( $p < 0.05$ for primary variants, $p < 0.0059$ for secondary variants)

Three significant associations were found between genetic variants and mucositis severity indicators after FDR correction (**Table 4**, **Supplementary Table S4**). *MTHFR* A1298C (rs1801131) was associated with lower odds of opioid use for oral mucositis (OR 0.34 [0.14–0.72], p=0.0035), while lower odds of TPN requirement were observed with miR-1206 rs2114358 (OR 0.09 [0.01– 0.43], p=0.0017) and *METTL3* rs1139130 (OR 0.08 [0.07–0.64], p=0.0020) (**Table 4**). 25 potential gene-drug interactions (p<0.2 for interaction term in the logistic regression model) were identified (**Table 5**). Interaction term p-values for all gene-drug interactions can be found in **Supplementary Table S5**.

**Table 4:** Significant Associations with Severity Indicators of Oral Mucositis.

| Severity Indicator | Allele | OR [95% CI] | p-value |
| --- | --- | --- | --- |
| Opioid use | *MTHFR* rs1801131 | 0.34 [0.14–0.72] | 0.0035* |
| TPN | miR-1206 rs2114358 | 0.09 [0.01–0.43] | 0.0017** |
| TPN | *METTL3* rs1139130 | 0.08 [0.07–0.64] | 0.0020** |

\* FDR-corrected p-value threshold 0.0059
\*\* FDR-corrected p-value threshold 0.0118

**Table 5:** Exploratory Gene-Drug Interactions with Oral Mucositis.

| **Drug** | **Variant** | **p-value** |
| --- | --- | --- |
| Actinomycin D | *ABCC4* rs7317112 | 0.0343 |
|  | *CAT* rs7943316 | 0.0881 |
|  | *METTL3* rs1139130 | 0.0633 |
|  | miR-4268 rs4674470 | 0.1069 |
| Anthracyclines | *ABCB1* rs1045642 | 0.0300 |
|  | miR-1206 rs2114358 | 0.1055 |
|  | *VDR* rs1544410 | 0.0435 |
| Alkylating agents<br>(cyclophosphamide<br>or ifosfamide) | *MTHFR* rs1801133* | 0.1134 |
|  | *CAT* rs7943316 | 0.0955 |
|  | *SLC19A1* rs2838958 | 0.0590 |
|  | *SLC19A1* rs1051266 | 0.1619 |
| Etoposide | *ABCB1* rs1128503 | 0.0990 |
|  | miR-1206 rs2114358 | 0.0571 |
|  | *SLCO1B1* rs2306283 | 0.1261 |
|  | miR-3683 rs6977967 | 0.0604 |
|  | *TYMS* rs34743033 | 0.1779 |
| Platinum<br>compounds<br>(carboplatin or<br>cisplatin) | *MTHFR* rs1801133* | 0.0907 |
|  | *SLCO1B1* rs2306283 | 0.1252 |
|  | *CYP3A5* (non-*1 alleles) | 0.1658 |
| Methotrexate | *ABCB1* rs1045642* | 0.1372 |
|  | miR-1206 rs2114358 | 0.0172 |
|  | miR-3683 rs6977967 | 0.1106 |
|  | *SLCO1B1* rs2306283 | 0.1986 |
|  | *CAT* rs7943316 | 0.0576 |
|  | *SLC19A1* rs2838958 | 0.1444 |

## 4.0 Discussion

Our study is one of few investigating genetic associations with oral mucositis in patients with solid tumours. We identified *MTHFR* A1298C (rs1801131) as having a protective effect against oral mucositis; each additional C allele was associated with a 65% lower odds of NCI-CTCAE grade ≥2 oral mucositis. This variant was also associated with a lower peak grade of oral mucositis and lower odds of opioid use for mucositis. Neither of our pre-specified variants of interest (*MTHFR* rs1801133 and *ABCB1* rs1045642) demonstrated significant associations with oral mucositis.

*MTHFR* A1298C (rs1801131) causes a glutamate-to-alanine substitution, consequently reducing methylenetetrahydrofolate reductase (MTHFR) activity to approximately 68% of the wild-type (compared to 45% in rs1801133).^20^ MTHFR catalyses conversion of 5,10-methylenetetrahydrofolate to 5-methyltetrahydrofolate, which plays an important role in DNA/RNA methylation pathways.^21^ One potential mechanistic explanation of our observed protective effect is that greater levels of 5,10-methylenetetrahydrofolate (due to reduced conversion) may allow for greater thymidylate synthesis, which is necessary for DNA synthesis in rapidly dividing oral mucosal cells. However, the impact of MTHFR on folate metabolism is complex and multifactorial, and as such this proposed mechanism remains speculative.

Previous literature studying the effects of *MTHFR* A1298C (rs1801131) on oral mucositis has yielded inconsistent results. One candidate gene study, performed in adults receiving haematopoietic stem cell transplant for chronic myeloid leukaemia, showed that copies of the C allele were associated with lower severity of oral mucositis.^22^ This finding is directionally consistent with that of our study. In contrast, a candidate gene study of 150 children with acute lymphoblastic leukaemia reported that patients with an AC or CC genotype (i.e. dominant allele model) had a significantly increased risk of oral mucositis (p=0.007).^23^ Several other studies did not find any association between *MTHFR* A1298C genotype and incidence of oral mucositis.^17, 24–29^

This variation in outcomes may be attributed to several factors, including differences in study populations, toxicity endpoints, and analytical methods. The most pertinent difference is that most of these cohorts were exposed to methotrexate, which interferes with folate metabolism and thus may influence genotype-toxicity associations. Meanwhile, only 12 (11.9%) of patients in our cohort received methotrexate. Similarly, the majority of these studies investigated solely haematological malignancies, while our cohort was exposed to a more diverse range of mucotoxic chemotherapy drugs. Ethnicity was also a key difference; while our population consisted of an Australian population with diverse self-reported ancestry, comparable studies were more focused on narrow geographical groups (e.g. Chinese, Indian, Slovenian, Egyptian, and Dutch populations). Notably, we did not utilise a genetic ancestry-informed approach to our analysis and thus cannot accurately comment on the ancestry proportions within our cohort. Mucositis toxicity endpoints were also heterogeneous; only den Hoed et al. specified use of the NCI-CTCAE criteria as determining occurrence of oral mucositis.^29^ Our toxicity endpoint was NCI-CTCAE grade ≥2 (i.e. modified diet or reduced oral intake), allowing clearer delineation between mucositis and non-mucositis patients. Lastly, an additive genetic model was only used by Robien et al.; Roy Moulik et al. and den Hoed et al. both used dominant genetic models, and the remaining studies used categorical analyses.^22, 23, 29^ This significant heterogeneity limits comparison between our findings to the existing literature and may explain our discordant findings.

Another important consideration is the likely polygenic and multifactorial nature of oral mucositis. Multiple pathways may plausibly cause or influence mucositis, affecting each of the distinct stages of its pathogenesis. Phenotype is also likely influenced by external treatment-related factors such as chemotherapy type, cumulative exposure, supportive care (including prophylaxis). This is particularly the case for a heterogeneous population such as ours, where such factors may have obscured the impact of any single genetic variant.

We also found significant associations between two genetic variants (miR-1206 rs2114358 and *METTL3* rs1139130) and requirement for TPN secondary to oral mucositis. However, neither variant was directly associated with grade ≥2 oral mucositis (OR 0.75 [0.38–1.44]; p=0.3805, and OR 0.73 [0.39–1.32]; p=0.2939, respectively), and the event rate of TPN requirement was low in our cohort (n=5; 5.0%). Furthermore, TPN requirement may be influenced by a range of determinants beyond severity of oral mucositis, including poor enteral intake, nausea and vomiting, nutritional status, and clinician preference. These findings should therefore be interpreted cautiously and considered hypothesis-generating.

Our study also found 25 exploratory gene-drug interaction pairs at the pre-defined screening threshold of p<0.2, though further confirmation was limited by low sample size. The lowest interaction p-value observed was for miR-1206 rs2114358 and methotrexate exposure (interaction p=0.0172). This variant has also previously been specifically linked to methotrexate-induced oral mucositis.^30^ miR-1206 has been implicated in the regulation of genes involving methotrexate transport and pharmacodynamics, including *SLCO1A2*, *ABCC2*, *ABCG2*, *TYMS*, and *FPGS*.^31^ These may influence methotrexate influx and efflux, as well as folate and thymidylate pathways. Alteration in miR-1206 structure may thus affect expression of these genes, resulting in increased downstream susceptibility to methotrexate-induced oral mucositis. Another signal of interest was observed between *ABCB1* rs1045642 and anthracyclines (interaction p=0.0300). *ABCB1* encodes P-glycoprotein, an ATP-dependent drug efflux transporter for which anthracyclines are known substrates.^32^ Variation in *ABCB1* expression or function could therefore plausibly modify anthracycline-associated toxicity risk. These exploratory findings require validation in larger cohorts before conclusions can be drawn about their contribution to oral mucositis risk.

Our study has several strengths, particularly its focus on children with solid tumours. This is an underrepresented population in the previous literature, which has primarily focused on haematological malignancies. In contrast, solid tumours are considerably more heterogeneous, are treated with a broader range of different chemotherapy regimens but still retain a high burden of oral mucositis. Our study addresses this important gap within pharmacogenomic literature. Furthermore, our cohort consisted of an ancestrally diverse paediatric cohort, while previously published literature on this matter has mostly been limited to adult cohorts or single-ethnicity cohorts. We also used NCI-CTCAE grade ≥2 as our primary toxicity endpoint, as this is often more clinically meaningful and impactful on children’s quality of life. Our use of PCR-free sequencing is also a methodological strength, minimising GC bias and ensuring broad genome coverage and high variant call rates. Finally, our exploration of hypothesis-generating gene-drug interactions is also important as different chemotherapy drugs and classes carry different oral mucositis risk profiles. Investigating these may thus provide a more nuanced understanding of the genetic determinants of chemotherapy-induced oral mucositis in children.

The primary limitation of our study was its modest sample size. A priori power calculations indicated only 35% power to detect a moderate genetic effect (OR 2.0) for the primary variants of interest. This was particularly restrictive for sub-analyses with low event counts, such as TPN requirement, and multivariable analyses such as gene-drug interaction models. Furthermore, a candidate gene approach prevented us from identifying variants of interest outside our pre-defined genetic panel. Our genetic panel was limited to genetic variants that had previously demonstrated significant associations with oral mucositis, which may have introduced selection bias to our study design. We also did not utilise ancestry-specific analyses and thus could not determine whether our findings remained consistent across different ethnic groups, or whether certain alleles differed in frequency across different ethnic groups. While our genetic data were collected within a prospective study framework, clinical data on oral mucositis was collected retrospectively from the electronic medical record and may be susceptible to inaccurate classification due to variation in clinical documentation. This is particularly relevant as the NCI-CTCAE criteria relies on subjective judgement rather than objective findings. Lastly, we were unable to validate our findings in an independent population, limiting their clinical translation.

While our study has demonstrated interesting potential associations, particularly that of *MTHFR* rs1801131 with a protective effect against oral mucositis, further research is needed before these findings can be integrated into clinical practice. Validation of our findings in an independent cohort is required, as well as studies with larger sample sizes and an ancestry-informed approach. Future research should incorporate multifactorial models to evaluate polygenic and pathway-level contributions to oral mucositis susceptibility, and to determine whether clinical covariates interact with genetic variation to influence toxicity outcomes. Ultimately, a greater understanding of genetic determinants predisposing to oral mucositis may enable us to better manage and prevent this toxicity in this vulnerable patient population.

## 5.0 Conclusion

Our candidate pharmacogenomic study of oral mucositis is one of few to investigate children with solid tumours. The primary finding of this study is that *MTHFR* A1298C (rs1801131) was associated with lower odds and lower severity of chemotherapy-induced oral mucositis. A range of other pre-specified genetic variants, including *MTHFR* C677T (rs1801133) and *ABCB1* rs1045642, were not associated with oral mucositis. We also identified a number of exploratory gene-drug interactions, including miR-1206 rs2114358 with methotrexate and *ABCB1* rs1045642 with anthracycline exposure. However, our findings were limited by modest sample size and absence of independent validation and thus require cautious interpretation. Larger studies incorporating ancestry-informed analyses and multivariable modelling with clinical covariates are recommended to validate our findings and further clarify the role of genetic variation in susceptibility to oral mucositis in this population.

## Conflicts of Interest

FJR receives institutional and salary support as a) a coinvestigator and subcontractor with the Peter MacCallum Cancer Centre for an investigator-initiated trial which receives funding support from Regeneron Pharmaceuticals; and b) a co-investigator on a translational research project funded by a Regeneron Pharmaceuticals grant.

FJR received travel expenses by MGI Australia and New Zealand.

## Acknowledgements

The Novo Nordisk Foundation Center for Stem Cell Medicine, reNEW, is supported by a Novo Nordisk Foundation grant number NNF21CC0073729.

## Artificial Intelligence Statement

OpenAI ChatGPT 5.5 was used to assist with paraphrasing of selected text to improve the clarity and readability of our manuscript. Artificial intelligence was not used to generate scientific conclusions, analyse data, or introduce any new content to our manuscript. All AI-assisted text was reviewed and edited by the authors. Originality and accuracy of the manuscript text, tables, figures, and references have been verified by the authors, who take full responsibility for the integrity of this work.

## Data Availability Statement

The data that support the findings of this study are available from the corresponding author, RC, upon reasonable request.

## Author Contributions

AC: conceptualisation, formal analysis, investigation, methodology, project administration, visualisation, writing – original draft, writing – review and editing

AH: data curation, investigation, writing – review and editing

MS: data curation

AG: formal analysis, methodology, writing – review and editing

FR: supervision

CM: writing – review and editing

SC: writing – review and editing

RC: conceptualisation, supervision, writing – review and editing

## Abbreviations

AGRF: Australian Genome Research Facility
BED: Browser Extensible Data
DNA: Deoxyribonucleic acid
DRAGEN: Dynamic Read Analysis for GENomics
FDR: False Discovery Rate
HREC: Human Research Ethics Committee
HWE: Hardy-Weinberg Equilibrium
MAF: Minor Allele Frequency
MARVEL-PIC: Minimising Adverse Reactions and Verifying Economic Legitimacy – Pharmacogenomic Implementation in Children
MTHFR: Methylenetetrahydrofolate reductase
NCI-CTCAE: National Cancer Institute Common Terminology Criteria for Adverse Events OR Odds Ratio
PCR: Polymerase chain reaction
RCHM: Royal Children’s Hospital, Melbourne
REG: Research, Ethics & Governance
RNA: Ribonucleic acid
TPN: Total Parenteral Nutrition
VCF: Variant Call Format
VCGS: Victorian Clinical Genetics Services
WGS: Whole Genome Sequencing

