## Supplementary Table S2 for "Genetic Determinants of Chemotherapy-Induced Oral Mucositis in Children with Solid Malignancies"

**Supplementary Table S2: Sensitivity analyses of genetic associations with grade ≥2 oral mucositis under dominant and recessive models**

| **Variant** | **Dominant Model** | | **Recessive Model** | |
| --- | --- | --- | --- | --- |
|  | **OR [95% CI]** | **p-value** | **OR [95% CI]** | **p-value** |
| *MTHFR* rs1801131 | 0.37 [0.15–0.88] | 0.0246 | Not converged | |
| *MTHFR* rs1801133* | 1.65 [0.69–3.98] | 0.2606 | 1.70 [0.22–10.83] | 0.5788 |
| *ABCB1* rs1045642* | 0.88 [0.34–2.41] | 0.7889 | 1.41 [0.57–3.42] | 0.4477 |
| *ABCB1* rs1128503 | 0.55 [0.22–1.39] | 0.2005 | 1.28 [0.50–3.18] | 0.6025 |
| *ABCC2* rs717620 | 1.58 [0.61–3.99] | 0.3415 | Not converged | |
| *ABCC4* rs7317112 | 0.86 [0.36–2.05] | 0.7267 | 0.59 [0.09–2.56] | 0.5076 |
| *ABCG2* rs2231142 | 0.68 [0.22–1.83] | 0.4545 | Not converged | |
| miR-1206 rs2114358 | 1.44 [0.59–3.62] | 0.4246 | 1.49 [0.36–5.37] | 0.5604 |
| miR-3683 rs6977967 | 1.50 [0.61–3.66] | 0.3709 | 1.70 [0.22–10.83] | 0.5788 |
| *SLCO1B1* rs2306283 | 1.11 [0.45–2.90] | 0.8232 | 0.61 [0.16–1.88] | 0.4014 |
| *CAT* rs7943316 | 0.65 [0.27–1.59] | 0.3458 | 1.49 [0.36–5.37] | 0.5604 |
| *VDR* rs1544410 | 0.51 [0.17–1.55] | 0.2254 | 1.20 [0.47–2.96] | 0.7024 |
| *VDR* rs2228570 | 1.90 [0.79–4.79] | 0.1535 | 1.45 [0.45–4.28] | 0.5175 |
| *CYP3A5* (non-*1 alleles) | 0.39 [0.04–3.34] | 0.3603 | 0.74 [0.29–1.97] | 0.5390 |
| *METTL3* rs1139130 | 0.44 [0.16–1.23] | 0.1171 | 0.90 [0.34–2.24] | 0.8232 |
| *SLC19A1* rs2838958 | 0.87 [0.28–2.98] | 0.8081 | 1.35 [0.56–3.25] | 0.4973 |
| miR-4268 rs4674470 | 0.62 [0.25–1.50] | 0.2933 | 0.99 [0.14–4.92] | 0.9932 |
| *TYMS* rs34743033 | 1.15 [0.35–4.44] | 0.8278 | 0.86 [0.34–2.09] | 0.7382 |
| *SLC19A1* rs1051266 | 0.93 [0.39–2.24] | 0.8710 | 1.62 [0.50–4.88] | 0.4069 |

* primary variants

** significant (p<0.05 for primary variants, p<0.0059 for secondary variants)
