## Supplementary Table S3 for "Genetic Determinants of Chemotherapy-Induced Oral Mucositis in Children with Solid Malignancies"

**Supplementary Table S3: Relationship between genetic variants and peak mucositis grade**

| **Variant** | **OR [95% CI]** | **p-value** |
| --- | --- | --- |
| *MTHFR* rs1801131 | 0.31 [0.16–0.57] | <0.001 |
| *MTHFR* rs1801133* | 1.43 [0.73–2.78] | 0.291 |
| *ABCB1* rs1045642* | 1.00 [0.61–1.63] | 0.998 |
| *ABCB1* rs1128503 | 0.84 [0.51–1.38] | 0.491 |
| *ABCC2* rs717620 | 1.26 [0.53–2.92] | 0.601 |
| *ABCC4* rs7317112 | 0.95 [0.52–1.72] | 0.871 |
| *ABCG2* rs2231142 | 0.63 [0.26–1.41] | 0.281 |
| miR-1206 rs2114358 | 1.21 [0.66–2.22] | 0.542 |
| miR-3683 rs6977967 | 1.31 [0.67–2.50] | 0.423 |
| *SLCO1B1* rs2306283 | 0.82 [0.47–1.41] | 0.474 |
| *CAT* rs7943316 | 1.09 [0.59–2.03] | 0.772 |
| *VDR* rs1544410 | 1.18 [0.67–2.11] | 0.562 |
| *VDR* rs2228570 | 1.33 [0.80–2.20] | 0.267 |
| *CYP3A5* (non-*1 alleles) | 0.86 [0.37–2.05] | 0.730 |
| *METTL3* rs1139130 | 0.82 [0.48–1.41] | 0.474 |
| *SLC19A1* rs2838958 | 1.09 [0.64–1.88] | 0.742 |
| miR-4268 rs4674470 | 0.82 [0.43–1.51] | 0.533 |
| *TYMS* rs34743033 | 0.91 [0.52–1.58] | 0.732 |
| *SLC19A1* rs1051266 | 1.10 [0.65–1.84] | 0.722 |
