## Supplementary Table S4 for "Genetic Determinants of Chemotherapy-Induced Oral Mucositis in Children with Solid Malignancies"

**Supplementary Table S4: Relationship between genetic variants and mucositis indicators**

| **Variant** | **Opioid use** | | **TPN use** | | **Unplanned admissions** | |
| --- | --- | --- | --- | --- | --- | --- |
|  | **OR [95% CI]** | **p-value** | **OR [95% CI]** | **p-value** | **OR [95% CI]** | **p-value** |
| *MTHFR* rs1801131 | **0.34 [0.14–0.72]** | **0.0035** | 0.22 [0.01–1.12] | 0.0717 | 0.43 [0.10–1.33] | 0.1519 |
| *MTHFR* rs1801133* | 1.32 [0.61–2.77] | 0.4708 | 1.53 [0.32–5.96] | 0.5603 | 1.18 [0.32–3.65] | 0.7876 |
| *ABCB1* rs1045642* | 0.96 [0.54–1.73] | 0.8879 | 0.84 [0.25–2.79] | 0.7707 | 1.32 [0.52–3.71] | 0.5640 |
| *ABCB1* rs1128503 | 0.67 [0.36–1.20] | 0.1773 | 0.67 [0.18–2.20] | 0.5116 | 0.77 [0.28–1.97] | 0.5792 |
| *ABCC2* rs717620 | 2.15 [0.81–5.61] | 0.1224 | 1.80 [0.23–11.42] | 0.5434 | 0.86 [0.12–4.01] | 0.8563 |
| *ABCC4* rs7317112 | 0.87 [0.42–1.74] | 0.6997 | 1.44 [0.35–5.42] | 0.5893 | 2.34 [0.80–7.10] | 0.1177 |
| *ABCG2* rs2231142 | 1.02 [0.38–2.46] | 0.9740 | 0.68 [0.04–3.85] | 0.7061 | 0.88 [0.13–3.41] | 0.8682 |
| miR-1206 rs2114358 | 0.58 [0.28–1.15] | 0.1204 | **0.09 [0.01–0.43]** | **0.0017** | 0.65 [0.22–1.98] | 0.4431 |
| miR-3683 rs6977967 | 1.58 [0.74–3.32] | 0.2293 | 2.71 [0.69–10.19] | 0.1441 | 2.54 [0.84–7.55] | 0.0958 |
| *SLCO1B1* rs2306283 | 0.85 [0.44–1.61] | 0.6144 | 0.13 [0.01–0.75] | 0.0184 | 0.41 [0.11–1.24] | 0.1186 |
| *CAT* rs7943316 | 1.09 [0.54–2.20] | 0.8068 | 3.19 [0.80–14.68] | 0.1005 | 1.98 [0.65–6.24] | 0.2260 |
| *VDR* rs1544410 | 1.31 [0.67–2.62] | 0.4364 | 1.13 [0.30–4.68] | 0.8619 | 0.95 [0.33–2.83] | 0.9190 |
| *VDR* rs2228570 | 1.64 [0.90–3.05] | 0.1080 | 0.80 [0.18–2.67] | 0.7212 | 1.08 [0.38–2.80] | 0.8822 |
| *CYP3A5* (non-*1 alleles) | 0.57 [0.26–1.28] | 0.1693 | 0.45 [0.12–1.99] | 0.2591 | 0.55 [0.19–1.95] | 0.3248 |
| *METTL3* rs1139130 | 0.60 [0.31–1.13] | 0.1122 | **0.08 [0.00–0.45]** | **0.0020** | 0.78 [0.28–2.16] | 0.6285 |
| *SLC19A1* rs2838958 | 0.91 [0.48–1.73] | 0.7591 | 0.41 [0.10–1.44] | 0.1619 | 1.03 [0.37–3.07] | 0.9587 |
| miR-4268 rs4674470 | 0.95 [0.44–1.94] | 0.8891 | 1.31 [0.28–4.83] | 0.7053 | 1.01 [0.28–3.01] | 0.9813 |
| *TYMS* rs34743033 | 0.76 [0.39–1.46] | 0.4021 | 0.62 [0.16–2.32] | 0.4673 | 1.45 [0.50–4.83] | 0.4998 |
| *SLC19A1* rs1051266 | 1.35 [0.72–2.51] | 0.3505 | 2.52 [0.74–9.80] | 0.1375 | 1.06 [0.36–2.82] | 0.9114 |
