## Supplementary Table S5 for "Genetic Determinants of Chemotherapy-Induced Oral Mucositis in Children with Solid Malignancies"

**Supplementary Table S5: Exploratory gene-drug interactions between genetic variants and mucotoxic chemotherapy drugs**

| **Variant** | **p-value** | | | | | |
| --- | --- | --- | --- | --- | --- | --- |
|  | **Actinomycin D** | **Anthracyclines** | **Alkylating agents** | **Etoposide** | **Platinum agents** | **Methotrexate** |
| *MTHFR* rs1801131 | 0.7875 | Not converged | 0.3802 | 0.2672 | 0.5116 | 0.5106 |
| *MTHFR* rs1801133* | 0.3762 | 0.7386 | **0.1134** | 0.3866 | **0.0907** | 0.6280 |
| *ABCB1* rs1045642* | 0.7345 | **0.0300** | 0.9816 | 0.6309 | 0.9772 | **0.1372** |
| *ABCB1* rs1128503 | 0.5386 | 0.5654 | 0.4355 | **0.0990** | 0.4857 | 0.8474 |
| *ABCC2* rs717620 | 0.6362 | 0.7696 | 0.6238 | 0.5481 | 0.5385 | 0.9407 |
| *ABCC4* rs7317112 | **0.0343** | 0.7251 | 0.5020 | 0.8350 | 0.5995 | 0.7630 |
| *ABCG2* rs2231142 | 0.7287 | 0.8424 | 0.8929 | Not converged | 0.6355 | 0.5823 |
| miR-1206 rs2114358 | 0.3705 | **0.1055** | 0.6107 | **0.0571** | 0.5845 | **0.0172** |
| miR-3683 rs6977967 | 0.3034 | 0.9076 | 0.4725 | **0.0604** | 0.5442 | **0.1106** |
| *SLCO1B1* rs2306283 | 0.8676 | 0.3301 | 0.3960 | **0.1261** | **0.1252** | **0.1986** |
| *CAT* rs7943316 | **0.0881** | 0.6997 | **0.0955** | 0.2891 | 0.5904 | **0.0576** |
| *VDR* rs1544410 | 0.4426 | **0.0435** | 0.6742 | 0.3579 | 0.5767 | 0.3269 |
| *VDR* rs2228570 | 0.8940 | 0.4980 | 0.9871 | 0.3792 | 0.8464 | 0.8777 |
| *CYP3A5* (non-*1 alleles) | 0.7407 | 0.3805 | 0.5647 | 0.2245 | **0.1658** | 0.7047 |
| *METTL3* rs1139130 | **0.0633** | 0.9899 | 0.6963 | 0.5311 | 0.6014 | 0.3302 |
| *SLC19A1* rs2838958 | 0.9927 | 0.4535 | **0.0590** | 0.4801 | 0.4344 | **0.1444** |
| miR-4268 rs4674470 | **0.1069** | 0.5177 | 0.9747 | 0.9880 | 0.3734 | Not converged |
| *TYMS* rs34743033 | 0.5894 | 0.6926 | 0.4020 | **0.1779** | 0.8055 | 0.7483 |
| *SLC19A1* rs1051266 | 0.7149 | 0.6959 | **0.1619** | 0.4178 | 0.9751 | 0.4622 |
