## Supplementary Material S1 for "Genetic Determinants of Chemotherapy-Induced Oral Mucositis in Children with Solid Malignancies": Supplementary Material S1.rtf

Supplementary Material S1: BED file used for variant calling

chr1	11794418	11794419	rs1801131	
chr1	11796320	11796321	rs1801133
chr7 	87509328	87509329	rs1045642
chr7	87550284	87550285	rs1128503
chr10	99782820	99782821	rs717620
chr13	95271268	95271269	rs7317112
chr4	88131170	88131171	rs2231142
chr8	128008932	128008933	rs2114358
chr7	7067004	7067005	rs6977967
chr12	21176803	21176804	rs2306283
chr11	34438924	34438925	rs7943316
chr12	47846051	47846052	rs1544410
chr12	47879111	47879112	rs2228570
chr7	99672915	99672916	CYP3A5*3 (rs776746)
chr7	99665211	99665212	CYP3A5*6 (rs10264272)
chr7	99652769	99652771	CYP3A5*7 (rs41303343; indel)***
chr14	21499771	21499772	rs1139130
chr21	45528652	45528653	rs2838958
chr2	219906501	219906502	rs4674470
chr18	657656	657657	rs34743033***
chr21	45537879	45537880	rs1051266
